# Detecting behavioural bias in GPS location data collected by mobile applications

**DOI:** 10.1101/2023.11.06.23298140

**Authors:** Hamish Gibbs, Rosalind M. Eggo, James Cheshire

## Abstract

A commonly-used form of human mobility data, called in-app mobility data, is based on GPS locations collected from a panel of mobile applications. In this paper, we analysed daily travel patterns from in-app GPS data in the United Kingdom to identify characteristic modes of travel behaviour, and assessed whether certain behavioural modes were more common among users of different groups of mobile applications. We also explored the relative importance of different mobility behaviours for the topology of an aggregated travel network. Our findings point to the presence of behavioural bias in in-app mobility data driven by the interaction between mobile device users and specific mobile applications. Our study also presents a general methodology for detecting behavioural bias in in-app mobility data, allowing for greater transparency into the characteristics of in-app mobility datasets without risking individual privacy or identifying specific mobile applications underlying a given dataset. Overall, the analysis highlights the need to understand the process of data generation for in-app mobility data, and the way that this process can bias the collective dynamics reported in aggregate mobility data.

## 1. Introduction

Human mobility data derived from mobile phones are increasingly used to measure economic activity (1,2), predict the spread of disease (3–6), forecast travel demand (7,8), measure responses to natural disasters (9,10), and understand human social dynamics (11,12). There are a number of sources of mobility data used in these applications, ranging from Call Detail Record Data, which estimates mobility based on mobile phone connections to nearby cell towers, to GPS data, which is collected by GPS sensors in smartphone devices. One specific form of GPS mobility data, in-app mobility data, is collected from a panel of mobile applications using software called a software development kit (SDK) packaged inside of mobile applications installed on a smartphone (13). The use of in-app mobility data accelerated during the COVID-19 pandemic due to the increasing availability of publicly accessible mobility indices produced by major mobility data aggregation companies, and the pressing need to understand real-time patterns of activity related to lockdowns and travel restrictions (14,15). To minimise the risk of revealing personally disclosive information, granular mobility data from multiple applications are typically aggregated to describe population-level dynamics, ranging from measures of ‘activity’ at specific points-of-interest (16), to measures of travel volume between pairs of locations (8,17).

Aggregating individual-level mobility data removes information about the unique travel behaviours of individuals, as well as the identity of the mobile applications which collected individual-level location data. While this is important for preserving individual privacy, aggregation can also mask key biases inherent to the process of in-app mobility data generation. This leads to uncertainty about the specific travel behaviours that define collective dynamics recorded by a given dataset. A key unknown in the analysis of aggregated in-app mobility data is the degree to which these data measure a representative subset of individual behaviours, or are influenced by biased sampling of individuals or activities, due to bias introduced by the specific mobile application collecting location data. Applications for navigation, for example, could be activated only while an individual engages in certain activities (i.e. the morning commute), while other applications, like an entertainment application, could be activated during an individual’s periods of leisure, such as weekends or evenings.

The specific mobile applications comprising widely-used in-app mobility datasets from different providers is proprietary information which is rarely released to the public. However, there is some public information on the identity of mobile applications supporting widely used in-app mobility data products. Analysis of the terms of service of mobile applications listed on the Google Play App Store, for example, has shown that navigation, weather, and fantasy sports applications share location data with the UK-based data aggregator Huq, the provider of in-app mobility data on which this study is based (18). The activity-specific nature of some of these apps raises questions about the degree to which the composition of apps in an in-app mobility dataset drives the detection of specific modes of behaviour. The same concern applies to other large-scale in-app mobility datasets, such as the US-based data aggregator SafeGraph, which has collected data through a SDK from applications for navigation, weather, and prayer (19).

Because the identity of mobile apps in in-app mobility data is typically private, or removed through data transformations, there is currently little insight into how potential behavioural bias may affect existing in-app mobility datasets, and how differences in the behaviours detected by different mobile applications could influence the aggregate dynamics reported by these data. Because of the sensitive nature of individual location information, and the commercial sensitivity of the specific applications which collect a given in-app mobility dataset, there is a pressing need for methods to identify potential behavioural bias, without presenting risks to individual privacy or disclosing sensitive commercial information. In this analysis, we endeavour to provide a view “underneath” an aggregated travel network derived from in-app mobility data, seeking to understand whether the mobile applications used to collect the mobility data are biased towards the detection of specific modes of behaviour, as well as the impact that this bias has on the topology of the aggregate travel network.

Our analysis is based on an individual-level in-app mobility dataset collected from 45 mobile applications in 2019. This dataset includes an anonymized identifier for the mobile applications that collected each location observation. In this paper, we use clustering techniques to identify common “modes” of daily travel behaviour based on characteristics such as travel distance, predictability, and regularity of location visitation. We then assess the potential for app-based behavioural bias by comparing the relative proportion of behavioural modes detected by different groups of mobile applications. Finally, we assess the implications of the different modes of detected travel behaviours on mobility dynamics reported in an aggregate travel network created from the underlying individual-level mobility dataset. In addition to demonstrating potential behavioural bias in a specific large-scale in-app mobility dataset, this paper demonstrates a general approach to resolving questions about the quality and representativeness of in-app mobility data while maintaining individual privacy, and masking the identity of individual mobile applications which collected location data. This approach can strengthen future uses of in-app mobility data by increasing the transparency of the data generation process, without the disclosure of sensitive personal or commercial information, thereby strengthening subsequent analysis based on a given dataset.

## 2. Methods

### 2.1 Data description

We used GPS location data collected from a panel of 45 mobile applications between January 1^st^ 2018 and December 31^st^ 2019 supplied by ESRC Consumer Data Research Centre and collected by Huq Industries, a commercial location data aggregator in the United Kingdom. Unique device identifiers, and identifiers pertaining to the application that collected location data were anonymized prior to data sharing. Individual location histories were analysed within a secure data environment, with research outputs checked by data scientists prior to release to prevent the disclosure of personally identifiable information (20).

### 5.2 GPS data preprocessing

GPS observations in the original in-app dataset are collected from either the high-accuracy GPS sensor in the phone, or lower-accuracy A-GPS sensors, depending on the availability of GPS satellites (21). This leads to a bimodal distribution in the accuracy of location observations in the dataset (Supplemental Figure 1.1). We selected high-accuracy GPS observations in the original in-app mobility dataset based on an accuracy threshold of ≤ 100 metres, to ensure that location observations were collected by the high-accuracy GPS sensor in a mobile device (22). We then transformed sequences of GPS locations (recording the time and spatial coordinates of a device) into a diary of ‘stop points’ describing locations where a device remained within a defined spatial diameter 𝑑_1_ for a minimum time threshold 𝑡. We then used agglomerative clustering to reference nearby stop-points to the same location (defined by a distance radius 𝑑_2_) according to the method proposed in (23). This transforms a sequence of GPS location observations into a set of stop-points describing ‘visits’ to a set of locations for each device, with spatial coordinates, start- and end-times. The agglomerative clustering of near-by stop points allows for the detection of repeated visits to the same location over time. We chose to define a stop-point based on a spatial radius of 200m (𝑑_1_, 𝑑_2_) and a time duration of at least 5 minutes 𝑡. These parameters are based on established definitions of the minimum duration of activities in travel surveys, and average walking speed (22,24).

### 2.3 Sample selection

Due to turnover in the panel of users in the location dataset, individuals’ activity was recorded for varying durations. This is a well-documented limitation of mobile phone location data including in-app data of the type used in this study (25). We took two steps to account for this bias in the dataset: (1) we based our clustering of travel modes around ‘days’ of travel, rather than individual devices by splitting visits crossing midnight (00:00 GMT) into distinct visits (ending 23:59:59 and beginning 00:00:01); (2) We filtered the location dataset for days of high quality location sampling, defined as days with at least 300 minutes of recorded activity, based on previous work using similar mobile-phone in-app location data (26). We performed a further sensitivity analysis to understand the effect that this threshold had on the size of the sample used in subsequent analysis (see Supplemental Figure 1.2);

### 2.4 Defining characteristics of daily travel behaviour

To discriminate between different modes of daily travel, we computed eleven features based on the travel diaries of individual mobile devices (Table 1). We chose metrics which highlighted distance travelled (in total and as displacement from and identified home location); recurrent visitation to the same locations; predictability of a sequence of visited-locations; total number of locations visited per day; and the amount of time spent at home. We estimated the home location based on a simple heuristic which identifies the most frequently visited nighttime location (defined between 22:00 and 06:00) per month.

**Table 1.** Measures of daily travel activity. Measures of daily travel activity computed for individual days of device activity.

| Characteristic | Measures | Units | Description |
| --- | --- | --- | --- |
| Total distance travelled | Daily total | Kilometre | Total daily distance travelled |
| Radius of gyration ($r_g$) | Daily | Kilometre | Distance travelled around a daily “centre of mass” |
| Distance from home | Daily maximum, minimum, mean | Kilometre | Distance travelled away from identified home location |
| Frequency of visitation ($p(i)_{norm}$) | Maximum, minimum, mean | | Likelihood to visit previously-visited locations on a given day (normalised by days observed) |
| Normalised entropy ($H_{norm}$) | Daily | | Predictability of visitation with respect to all possible locations visited by a device (normalised by days observed) |
| Number of visits | Daily | Count | Daily number of locations visited |
| Time spent at home | Daily | Seconds | Amount of time spent at identified home location |

Some daily metrics, like the total distance travelled by a device, are directly comparable between devices, while frequency measures are related to the total amount of time in which a device is included in the dataset. Similarly, entropy is related to the total number of locations ever visited by an individual. To account for this dependence, we normalised frequency and entropy measures by the observation window of a device (defined as the number of days the device was observed), and the total number of locations visited by a device, respectively.

Descriptions of individual behaviour are based on a set of 𝑁 identified stop points 𝑉_1…*N*_ for each day of travel activity for each device. The sequence of stop points for a specific day are defined by stop point identifiers *Stop*_1,…*N*_, spatial coordinates (𝑥_1_, 𝑦_1_), (𝑥_2_, 𝑦_2_), . . . (𝑥_*N*_, 𝑦_*N*_), and time intervals 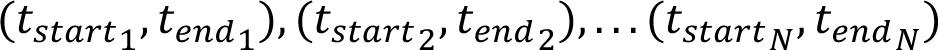.

We calculate distances (total distance travelled and distance from home) as the Euclidean distance between the locations of relevant visits.

Radius of gyration 𝑟_*g*_ is based on the centre of mass (𝑥_*cm*_, 𝑦_*cm*_) of a daily trajectory where:

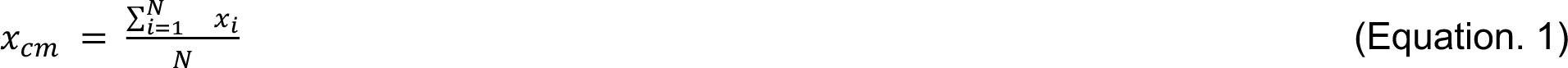

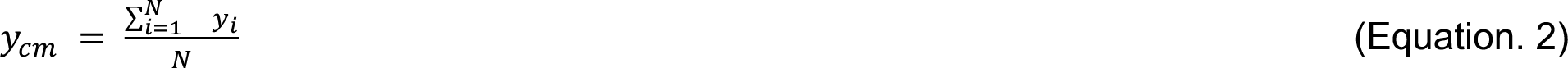

𝑟_#_ is derived from squared displacements 𝑟^2^_*i*_ from the centre of mass for each location 𝑖 in the trajectory:

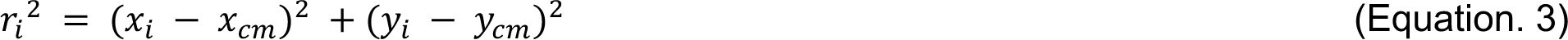

Thus:

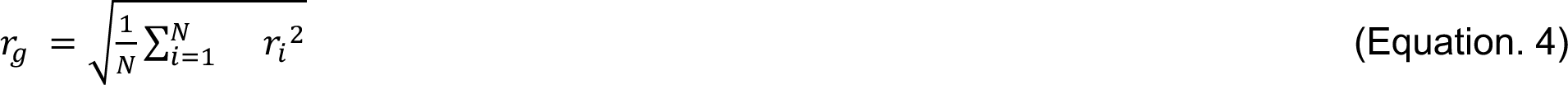

We calculate Normalised Frequency of Visitation 𝑝(𝑖)_$%&’_ for each location 𝑖 in the trajectory relative to the number of observation days for each device 𝐷 based on the number of times the location was visited by an individual 𝑣_3_.

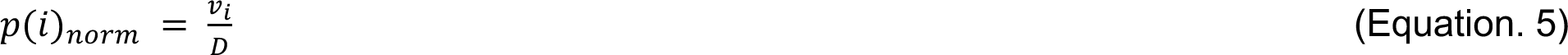

We calculate Normalised Entropy 𝐻_$%&’_ based on the probability of visiting individual location 𝑝(𝑖) based on the number of visits to each location 𝑣_3_ and the total number of visits to all locations 𝑉:

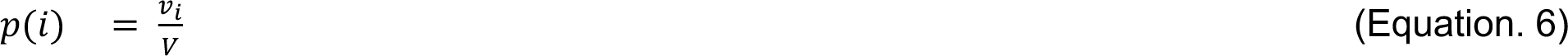

Entropy 𝐻 is then defined as:

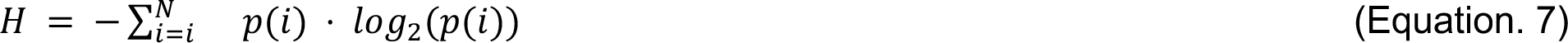

And Normalised Entropy 𝐻_$%&’_ is defined as:

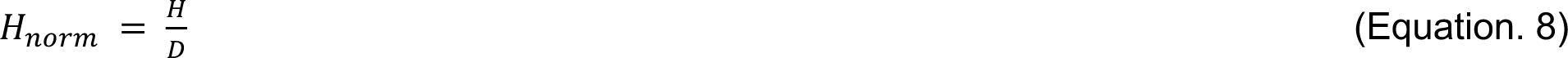

Number of Visits is the count of distinct visits recorded for an individual per-day, and Time Spent Home measures the total number of seconds an individual was observed in at their identified home location.

We used principal component analysis (PCA) to account for collinearity among the daily travel measures which can influence the results of clustering analysis, and to reduce the dimensionality of the daily travel measures, increasing the computational efficiency of the clustering. We assessed the correlation between input features prior to performing the PCA to understand possible axes of differentiation in the original data and to identify highly correlated input features (Supplemental Figure 2.1). After performing PCA, we assessed the correspondence between principal components and the original features by computing the Pearson Correlation Coefficient between the components and the original features (Supplemental Figure 2.2). This indicates the potential axes defining subsequent clustering of similar patterns of mobility activity. We used the first 6 principal components for subsequent analysis (a 45% dimensionality reduction) which captured >95% of the variance of the original dataset (Supplemental Figure 2.3).

### 2.5 Detecting modes of daily travel behaviour

We clustered the resulting principal components using the k-means algorithm (27). K-means is a widely used clustering algorithm which partitions a series of data points into pre-defined 𝑘 groups by repeated random selection of cluster centroids to identify the partition which minimises within-cluster variance. We chose the k-means algorithm due to its efficient implementation and ability to handle the clustering of large datasets (28). Our choice of clustering algorithm, combined with dimensionality reduction from the PCA allows for the efficient clustering of >10 million daily activity patterns.

To gain an intuitive understanding of the effect of different numbers of clusters on our dataset, we created a clustergram (29) for values of 𝑘 between 2 and 9. The clustergram displays the size, and PCA-weighted similarity of clusters as well as the transition of data between clusters at different values of 𝑘. This visualisation can be a useful tool to spot regularities in the dataset across different numbers of clusters (for example, “branches” of similar clusters at higher values of 𝑘). To choose an appropriate value of 𝑘, we then assessed the quality of the identified clusters using three measures: Silhouette Score (30), Davies-Bouldin Index (31), and Calinski-Harabasz Index (Variance Criterion Ration) (32). Each metric highlights different aspects of the clusters: Silhouette Score compares the within-cluster distance to the distance between the next-most-similar cluster, Davies-Bouldin Index measures the average of the maximum ratio of within-cluster distance to between-cluster distance for each cluster, and Calinski-Harabasz Index measures the ratio of the sum of within-cluster variance to between-cluster variance. We selected 4 clusters based on the value of *k* producing high Silhouette Score, low Davies-Bouldin Index, and high Calinski-Harabasz Index (Supplemental Figure 3.1).

Calculation of clustering quality metrics requires the computation of distances between all observations which is not practical for the size of our dataset, as the number of required distance calculations scales as 𝑂(𝑛^2^) to the number of observations (quadratic complexity). To reduce the number of required distance calculations, we performed bootstrapping for 1000 samples of 0.1% of the original dataset to produce a density estimate of each clustering quality metric.

### 2.6 Detecting application-based behavioural bias

We detect behavioural bias related to the mobile application that collected the location data by clustering mobile applications together based on the relative proportion of each behavioural mode identified by individual mobile applications. Similar to aggregating daily travel histories into behavioural modes to preserve individual privacy, we group applications into eight groups using k-means clustering, informed by the relative proportion of each behavioural mode identified by each individual mobile application. We then computed the distribution of stop points and total travel distance for daily travel histories recorded by each application group.

### 2.7 Effect on travel network connectivity

To assess the influence of identified modes of behaviour on the topology of the overall travel network and the implications of different behaviours detected by different groups of mobile applications, we constructed an aggregated travel network describing individual movements between cells in a hexagonal spatial grid with resolution equal to approximately 36 km^2^ (33). In this travel network, connections are defined by a pair of sequential stop-points recorded in different grid cells by an individual mobile device. Using this aggregated travel network, we then performed a series of experiments, removing days of travel corresponding to a specific behavioural mode from the aggregated network one-at-a-time. Finally, we assessed how this removal affected different measures of network topology: the total number of edges in the network, the average shortest path distance (both spatially-explicit and topological distance), the diameter (maximum distance between any nodes), and transitivity or clustering coefficient (describing the number of triangles or cycles of length three in the network).

## 3. Results

### 3.1 Detecting travel behaviour from in-app location histories

We analysed anonymized GPS location data recording travel by 782,000 mobile devices in England and Wales collected between January 1st, 2018 and December 31st, 2019. We filtered GPS positions for high accuracy observations (Supplemental Figure 1.1), and applied a two-stage ‘stop-point’ detection algorithm (23) to sequences of GPS locations which identified 43 million locations where mobile devices were stationary within a spatial radius of 200m for a minimum duration of 5 minutes (Figure 1a-d). We then selected days of travel with observation windows of at least 5 hours (300 minutes) per day (Supplemental Figure 1.2), yielding a dataset recording the positions of 10.9 million days of travel by 677,000 devices (Figure 1e).

**Figure 1.**
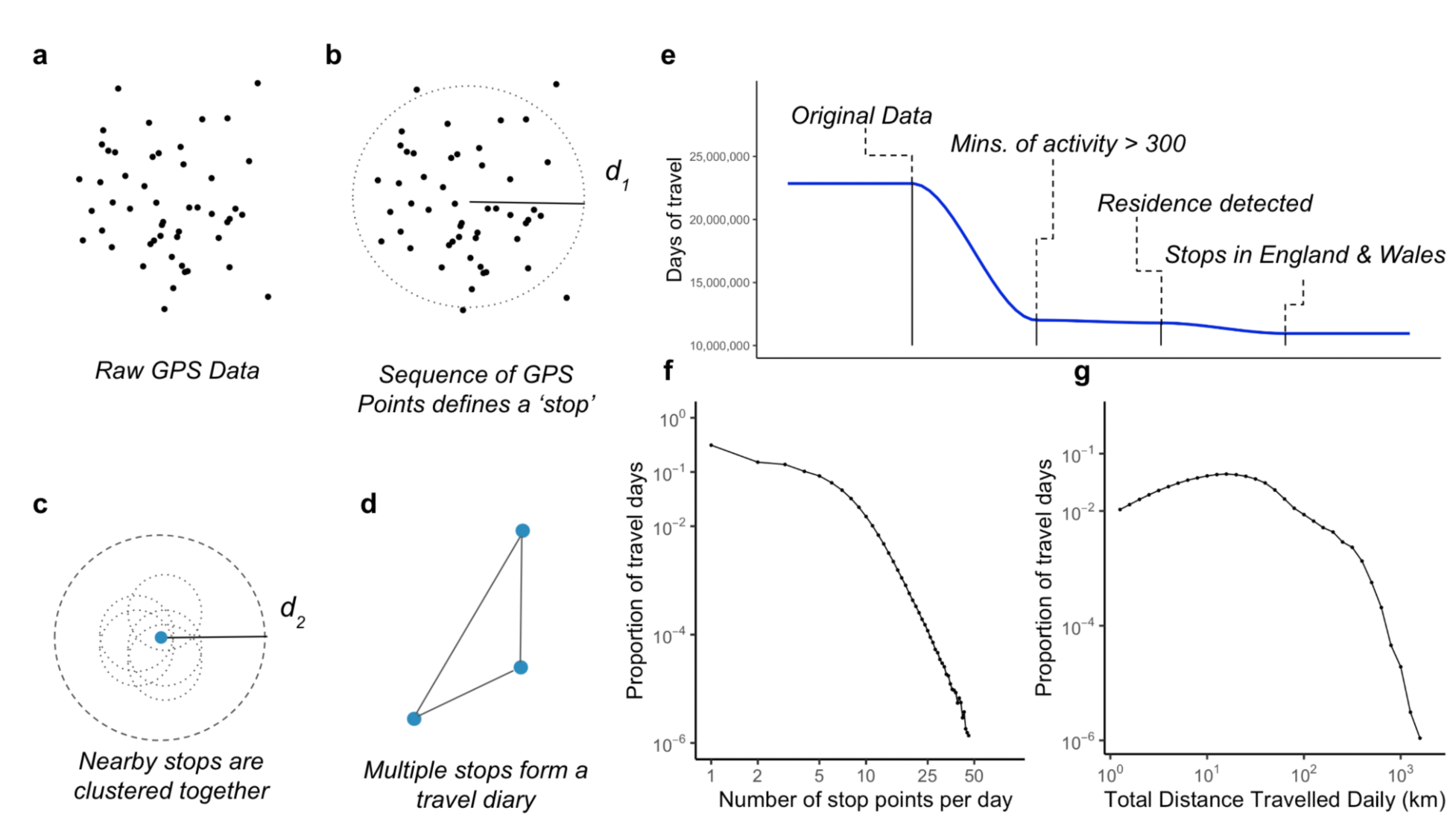
Overview of the mobility dataset. a-d) Diagrams illustrating the detection of stop-points from a sequence of GPS locations. Distance threshold d1 is the maximum spatial ‘roam’ of a device, d2 is the spatial threshold used to attribute nearby stops to the same location. e) The size of the mobile phone location dataset (total days of travel) through different stages of sample selection. f) The distribution of the number of stop-points for all days of travel and g) the total distance travelled for daily sequences of stop-points with travel greater than 1 km.

In the travel dataset, individual devices were present for varying length durations. Devices are recorded in the dataset for a median of 4 days (IQR: 1, 14). To account for the varying-length sampling windows, we frame our analysis around ‘days’ of travel as opposed to individual devices, where one device may contribute multiple days of travel to subsequent analysis. In the dataset, 31% of travel days recorded only one visited location, while 43% of travel days recorded a total distance travelled less than 1 km (Figure 1f,g).

### 3.2 Daily travel activity clustering

We identified modes of daily travel activity based on measures of travel distance, visitation frequency, and entropy (predictability) using k-means clustering informed by a principal component analysis of the characteristics of daily travel histories (Supplemental Figures 2.2, 2.3). The clustering analysis identified four clusters (Table 2): *regular-travel* (59% of travel days), *stay-at-home (38%* of travel days*), long-distance* (1.6% of travel days), and *away-from-home (0.9%* of travel days). We describe two of these clusters (*long-distance,* and *away-from-home*) as ‘long tailed’ clusters, referring to their relatively low frequency and the extreme distribution of some travel characteristics in these clusters, which are primarily defined by high total daily distance travelled (*long-distance)*, and high minimum daily distance from home (*away-from-home*) (Figure 2a). The distribution of total distance travelled (Figure 2b) highlights the difference in travel behaviour across clusters, with the *long-distance* cluster showing high total distance travelled, while *away-from-home* is distinguished by a high minimum distance travelled from home (Figure 2c).

**Table 2.** Description of identified modes of travel. The result of cluster analysis of travel based on distance, visit frequency, and entropy of daily travel patterns.

| Cluster | Descriptive title | Travel days | Description |
| --- | --- | --- | --- |
| 1 | *Regular-travel* | 59% | Variable total travel distance, low distance from home, higher likelihood to visit a new location, variable entropy |
| 2 | *Stay-at-home* | 38% | Low travel distance, low likelihood of visiting a new location, low entropy, high time spent at home |
| 3 | *Long-distance* | 1.6% | High distance travelled, low minimum distance from home, high likelihood of visiting new locations |
| 4 | Away-from-home | 0.9% | High minimum distance from home (never near home location), variable total distance, high likelihood of visiting a new location |

**Figure 2.**
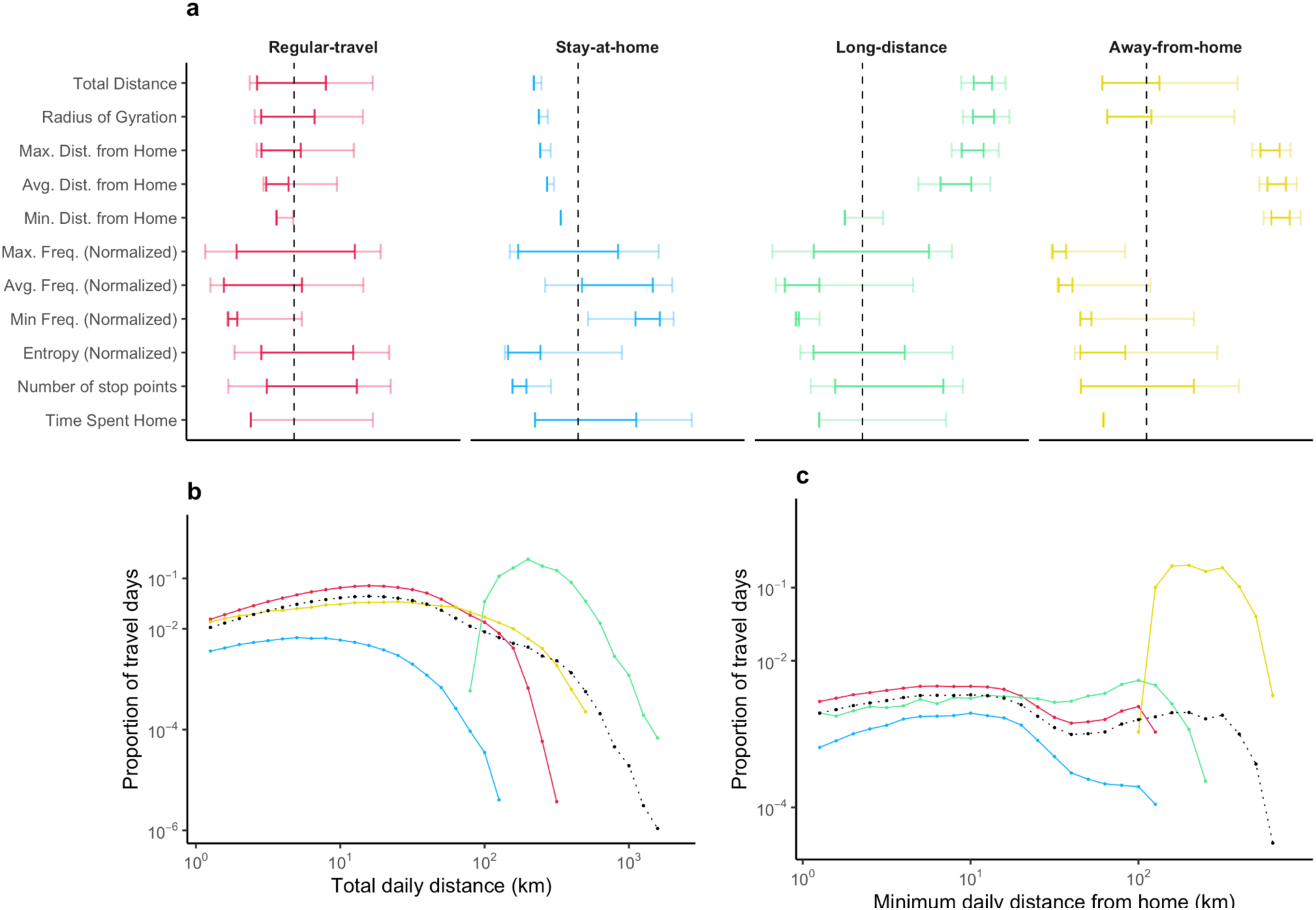
Clustering identifies long-distance, atypical travel. a) The distribution of travel measures for each identified cluster. Intervals indicate 90% and 50% density intervals for observations within each cluster. Values have been transformed for comparison across measurement scales. Dotted black line indicates average for each variable. b) The distribution of total distance travelled for each cluster. Dotted black lines in panels b and c indicate the variable distribution for the overall dataset. c) The distribution of the minimum distance travelled from home for each cluster.

### 3.3 Application-specific behavioural bias

To understand whether bias towards specific modes of behaviour is introduced by data collection through different mobile applications, we perform a clustering analysis of mobile applications in the dataset, grouping apps based on the distribution behavioural modes identified. While our primary motivation for grouping applications together is to obscure the identity of any single app in the original dataset, the identified groups reveal notable differences in the modes of behaviour captured by different mobile applications (Figure 3a). We find that three groups (A, B, C) detect a high percentage of *stay-at-home* activity, and relatively low *away-from-home* and *long-distance* travel behaviour, while other groups, (D, E, F, H) detect a relatively higher percentage of *long-distance* and *away-from-home* travel relative to the dataset overall. Only three groups (D, F, G) detect a higher relative quantity of *regular-travel*. It is important to note that different mobile applications contribute different volumes of data to the dataset, resulting in an unbalanced sampling of behaviours by different apps (Figure 3b). App groups D and G, for example, represent 13 apps contributing 828,000 days of travel, equal to 75% of the total days of travel recorded by the dataset (Figure 3b). This uneven contribution of different mobile applications increases the potential for behavioural bias introduced by the outsized contribution of a small group of mobile applications which capture a particular distribution of behavioural modes.

**Figure 3.**
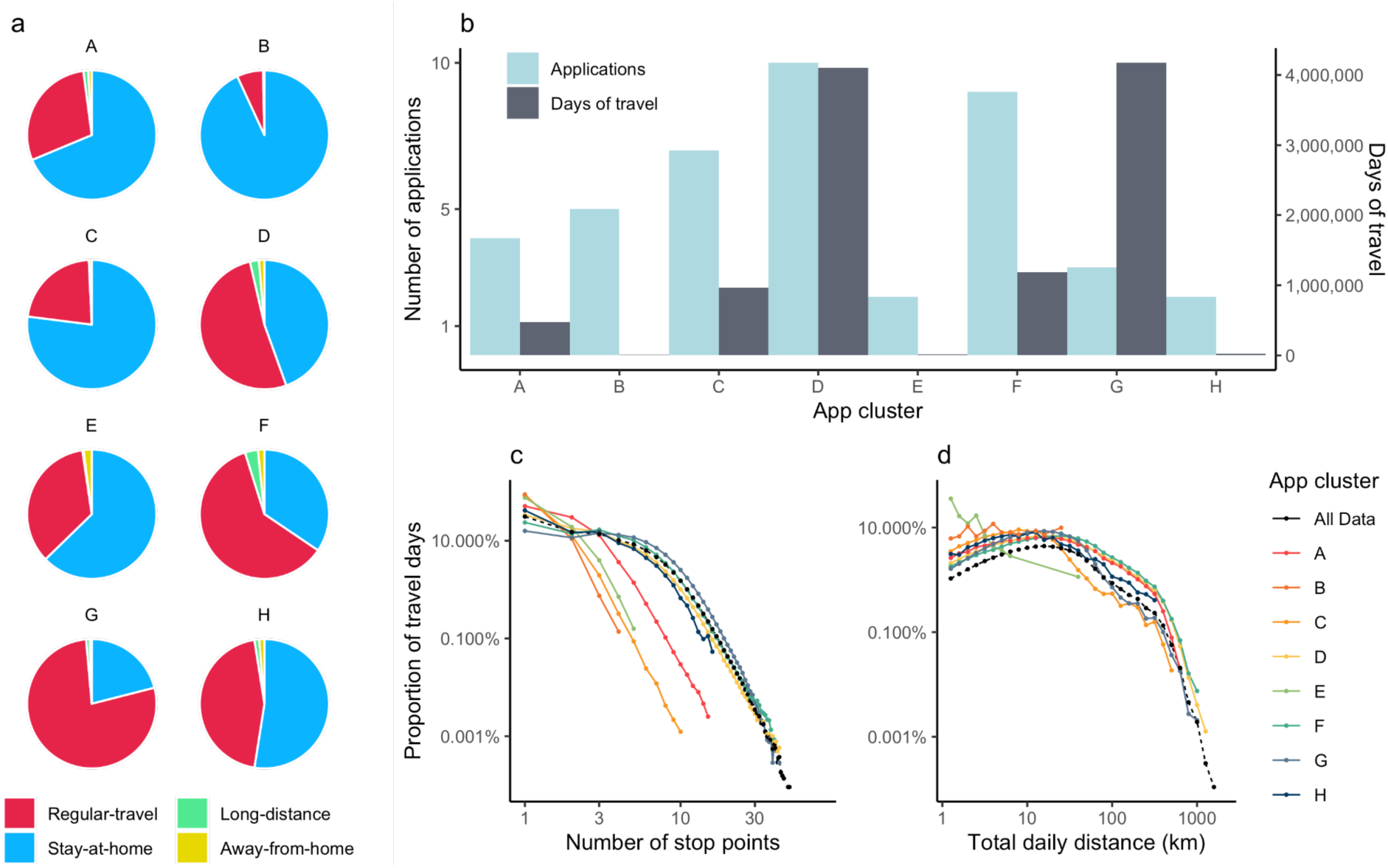
Distribution of behavioural modes detected by different mobile applications. a) Pie charts showing the distribution of behavioural clusters for each app cluster. b) Number of apps and days of travel in each app cluster. c) Distribution of visits for each app cluster. d) Distribution of total distance travelled for each app cluster.

The varying distribution of behavioural modes in each cluster produces different distributions of visits (Figure 3c) and daily travel distance (Figure 3d) for days of travel recorded by different groups of apps. Apps capturing high proportions of *stay-at-home* activity (A, B, C) have more days of travel with a low number of visits and shorter overall travel distance, compared to other app groups which report higher proportions of regular-travel, long-distance and away-from-home behaviours, with correspondingly higher volumes of travel days with high numbers of visits and longer distance travel. The daily total distance travelled for app group G shows an interesting artefact, with a notable drop in the frequency of days of travel with total distance above 50 km. This may reflect an outer limit to the total distance travelled per day by individuals recorded by apps in this group, due to limits such as the availability of transit or the mode of behaviour for users of these apps.

Analysis of the daily patterns of location data collection across different app groups roughly show three different motifs (Figure 4a). Some groups of apps (B, C, F) show low usage overnight and in the morning, with high usage during the daytime. Other app groups show a relatively consistent pattern of usage throughout the day (groups D, E, G, H) while one group (I), shows a peak usage after 8PM. Across days of the week (Figure 4b), the majority of app groups collect data primarily on weekdays, while one cluster (B) collects data primarily on weekends.

**Figure 4.**
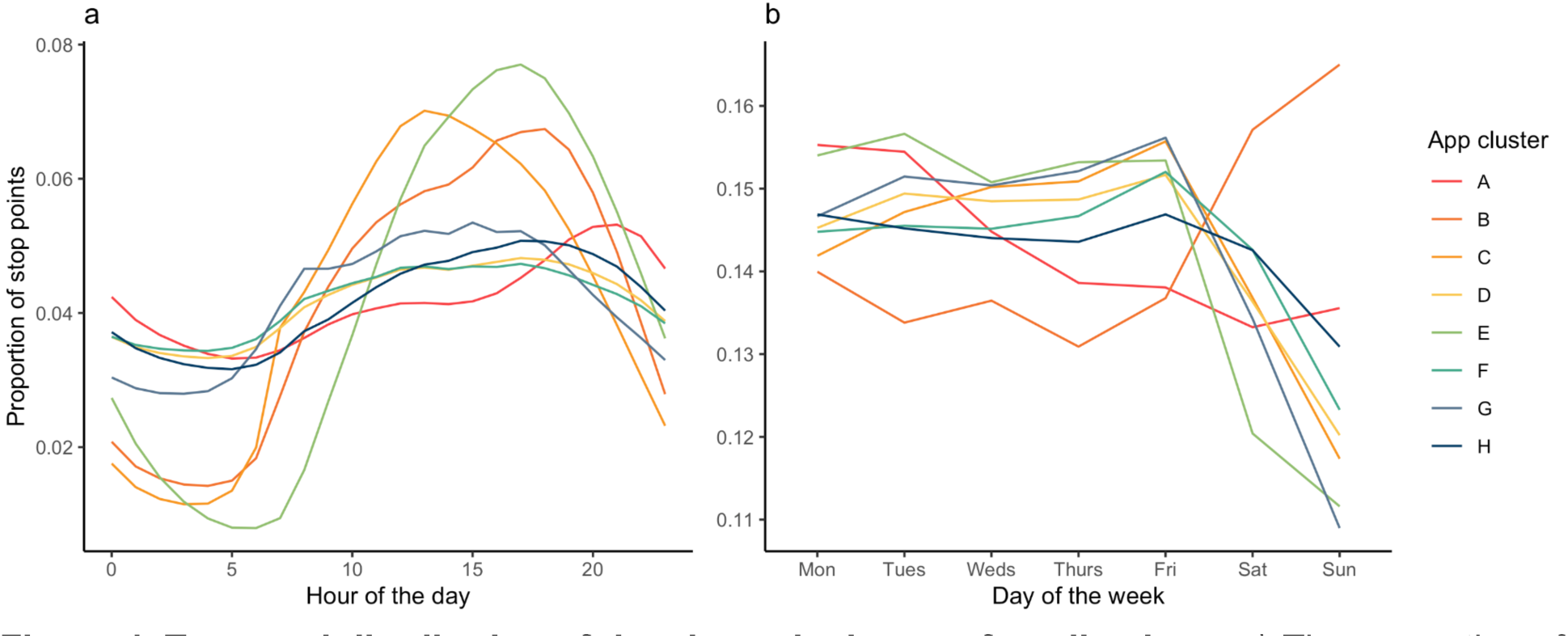
Temporal distribution of data in each cluster of applications. a) The proportion of stop points detected at different hours of the day and b) days of week for each group of mobile applications.

This points to the potential that different groups of applications are detecting different modes of behaviour related to work- or leisure-related activities over time.

### 3.4 Behavioural clusters drive travel network connectivity

To understand how the identified modes of travel behaviour influence the topology of the travel network, and the potential implication of different modes of behaviour collected by different mobile applications, we aggregated movements into origin-destination flows across a regular spatial grid of approximately 36 km^2^ (33). We then performed an experiment, removing one behavioural mode at a time, to observe the resulting changes on the travel network topology (Table 3). We found that *regular-travel* formed 42% of the unique network connections (‘edges’) in the aggregated network, and that *long-distance* movements, although they comprised 1.6% of all travel days, represented 35% of the unique edges in the network. By contrast, the other ‘tail’ cluster, *away-from-home,* represented only 1.9% of unique edges, while removal of *stay-at-home* movements had almost no influence on the remaining travel network.

**Table 3.**
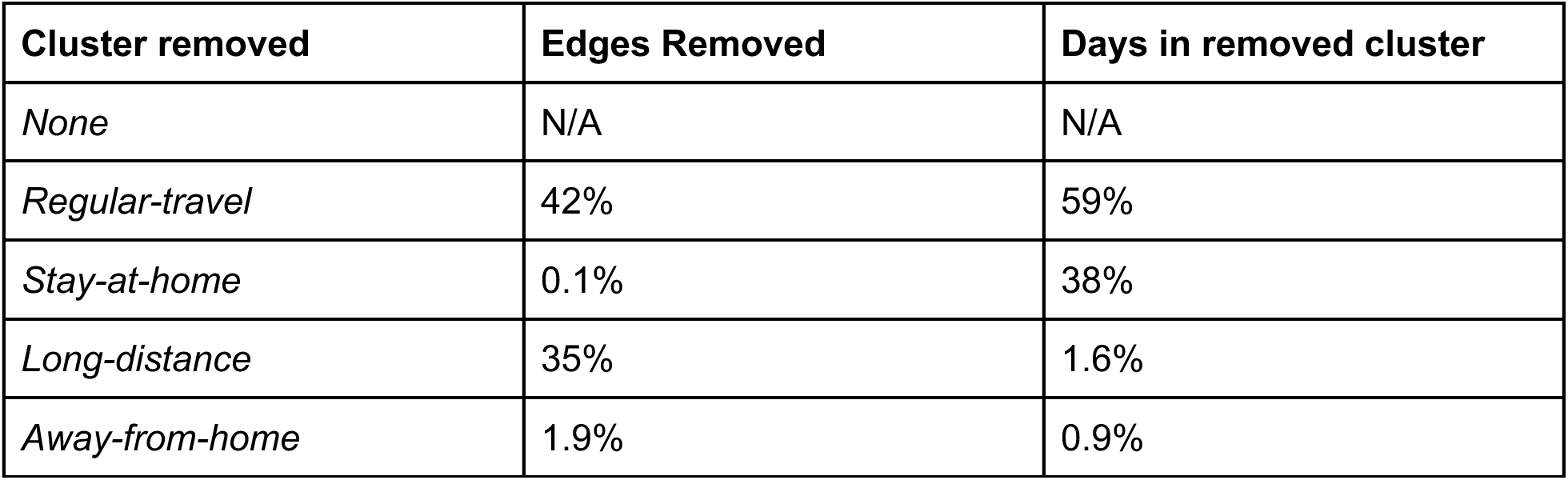
Number of edges removed by each cluster compared to the proportion of travel days. Shows changes in the number of edges in the aggregated travel network caused by the removal of different clusters of daily travel activities.

| Cluster removed | Edges Removed | Days in removed cluster |
| --- | --- | --- |
| *None* | N/A | N/A |
| *Regular-travel* | 42% | 59% |
| *Stay-at-home* | 0.1% | 38% |
| *Long-distance* | 35% | 1.6% |
| *Away-from-home* | 1.9% | 0.9% |

The number of unique edges represented by each cluster is only one means of assessing changes to the topology of the travel network. We computed additional measures to better illustrate how the aggregated travel network responded to the removal of each cluster of travel behaviours (Table 4). We found that some spatially explicit measures, such as the diameter of the network (maximum distance between any nodes), and the spatially-explicit mean shortest path length, remained similar with any cluster removed. This is because the network retained the same spatial extent despite the removal of some movement modes. The topology of the network, however, showed sensitivity to the removal of certain clusters of behaviour, in particular to *long-distance* movements.

**Table 4.** Measures of network characteristics with different behavioural clusters removed. A collection of measures characterising the spatial and topological changes in the travel network with the removal of different activity clusters.

| Cluster removed | Diameter (km) | Mean shortest path (km) | Mean shortest path (Topological) | Transitivity |
| --- | --- | --- | --- | --- |
| *None* | 750.79 | 228.58 | 2.62 | 0.26 |
| *Regular-travel* | 1048.40 | 231.65 | 2.75 | 0.20 |
| *Stay-at-home* | 760.27 | 228.60 | 2.62 | 0.26 |
| *Long-distance* | 763.40 | 229.09 | 3.56 | 0.38 |
| *Away-from-home* | 753.45 | 228.67 | 2.62 | 0.26 |

Changes in the degree and edge-distance distributions with the removal of a specific mode of behaviour provide further insight into the topological changes to the travel network. Decreases in high degree nodes (observed with the removal of *regular-travel* and *long-distance* modes) indicate a network with fewer well-connected locations (Figure 5a). Decreases in the edge-distance distribution (Figure 5b) indicate fewer long-distance connections in the network. For both measures, removal of the *stay-at-home* and *away-from-home* modes produced travel networks with characteristics similar to the original dataset. It is important to note that differences in travel network topology are sensitive to the spatial scale at which individual movements are aggregated. We perform a sensitivity analysis showing how different measures of travel network topology change with decreasing spatial resolution (Supplemental Section 4).

**Figure 5.**
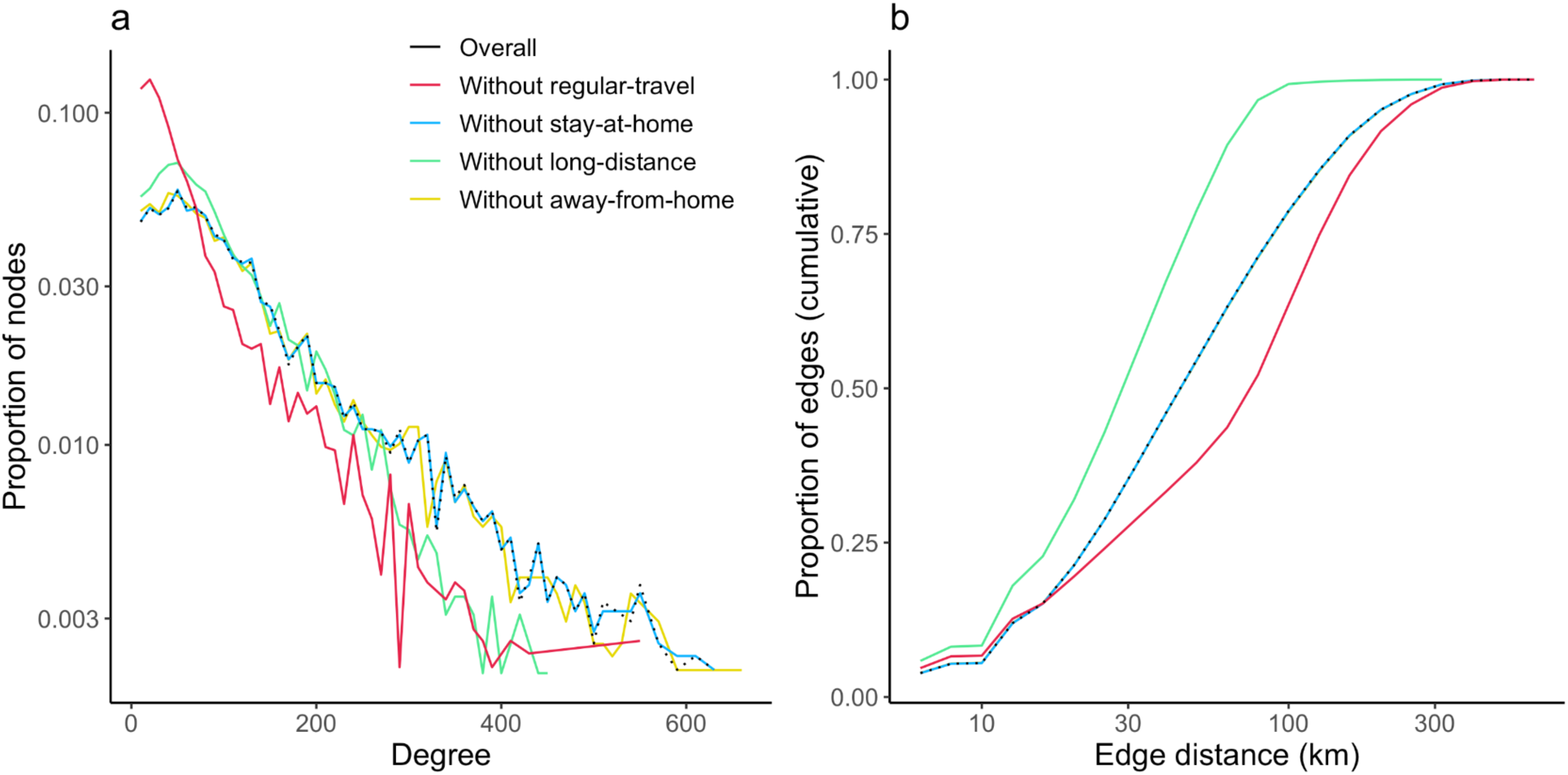
Degree and edge distance distribution with behavioural clusters removed. a) The degree distribution and b) edge distance distribution for the aggregated travel network, for travel networks with each activity cluster removed one-at-a-time.

## 4. Discussion

This analysis demonstrates that mobile applications underlying a large-scale in-app mobility dataset introduce bias towards specific travel behaviours through differential measurement of certain forms of travel activity. Moreover, the degree of this bias, and the behaviours which are over-or under-represented, is dependent on the group of applications which comprise the dataset. This confirms that the nature of mobility patterns recorded by a given source of in-app mobility data are intimately connected to the mobile applications from which the data was collected.

Aside from the over- or under-representation of certain modes of mobility behaviour, we also show how specific behavioural modes (like *long-distance* travel) can themselves have an outsized influence on the overall structure of collective dynamics recorded in an aggregated mobility dataset. In our analysis, for example, the *long-distance* mode, which accounts for 1.6% of travel days, forms 35% of the unique origin-destination pairs in an aggregate travel network. The combination of these two findings: that groups of mobile applications detect modes of mobility activity at different rates, and that certain behavioural modes play an outsized role in defining population-level measures of travel behaviour, raise questions about how to best account of the application-based sampling frame inherent to in-app mobility data in order to improve the robustness of conclusions drawn from these data.

In-app mobility data are an undeniably valuable form of mobile phone location data that have been used to address a range of scientific and practical challenges, in particular during the response to the COVID-19 pandemic. Unlike other forms of mobile phone mobility data, namely Call Detail Record data, in-app GPS data provide higher spatial granularity which can provide more detailed insights regarding patterns of social contact and visitation to points of interest (13). The challenge of behavioural bias in in-app mobility data, as explored in this study, is likely inherent to the structure of the data generation process which create in-app mobility data, and must be considered in addition to other well-known biases in large-scale mobile phone location data such as “drift” in the sample of individuals contributing location data over time (34), and concerns about bias in mobility data with respect to specific demographic groups (35). Moreover, although we have identified specific biases present in the dataset used in this study, it is likely that the composition of applications generating other sources of in-app mobility data will have their own unique patterns of behavioural bias.

The first step towards addressing application-based behavioural bias in in-app mobility data is to understand the extent to which different activity patterns detected in the dataset are attributable to different mobile applications. The methodology presented in this study provides a generalizable approach to answering this question, while maintaining the private character of both the individuals and mobile applications included in the underlying dataset. There are further opportunities to use this information to inform statistical re-weighting of in-app mobility data, by increasing or suppressing the contributions of specific mobile applications through post-stratification. Similar techniques are already used to reduce the effect of demographic bias in in-app mobility data (26,36,37). The challenge of applying post-stratification techniques, however, arises from the scarcity of representative information on the behavioural characteristics in a given population of interest. As, in-app mobility data are often employed in domains for which there are few other potential sources of information, defining the correct distribution of behavioural modes for a given population may remain a challenge. Developing techniques in the generation of synthetic mobility data could also contribute to improving the accuracy of in-app mobility data, as synthetic data could “fill-in” missing data for specific modes of behaviour (38).

Ultimately, future uses of in-app mobility data can be strengthened by an improved understanding of the process of data generation underlying a given class of mobility data (such as in-app data), and through rigorous exploration of the unique characteristics of specific mobility datasets. Because of the imperative to preserve individual privacy, and the ecosystem for processing and sharing mobility data, information on the quality of location datasets can often be lost through privacy-preserving data transformations prior to data sharing with researchers. In this analysis, we present a ‘middle path’ between full dataset transparency and a naive approach to dataset aggregation, in which we illuminate some of the behavioural and application-specific differences that drive important variations in the structure of recorded mobility data, while avoiding the risk of disclosing potentially sensitive information. This approach required the processing of sensitive individual-level mobile location data within a secure research facility, with adequate safeguards prior to exporting the specific insights presented in this study (20). In the future, the type of analysis presented here could be performed by location data providers, in order to improve transparency in the quality of an in-app mobility dataset, and inform the use of aggregated mobility data, such as aggregated travel networks, in subsequent analysis.

## 5. Conclusion

Our analysis highlights the need for scientific understanding of the behavioural biases present in sources of in-app mobility data. This understanding cannot be achieved merely through the analysis of aggregated mobility data, and highlights the tension between the imperative to preserve privacy, and the need to understand limitations within mobility data in order to improve the robustness of conclusions drawn from these data. Publicly available, aggregated mobility datasets could be accompanied by findings similar to the analysis presented in this paper, which can highlight the unique biases affecting a given dataset, and point to potential remedies in order to improve confidence in subsequent uses of the data.

## Competing Interests

The authors declare no competing interests.

## Ethics

This research has been approved by the University College London Research Ethics Service (ref: 21813/001).

## Data Availability

This analysis is based on GPS location data collected by the UK-based data aggregator Huq. Data is available for academic research by application to Huq Industries (https://huq.io/). Analysis of individual-level GPS trajectories was performed in a secure computing environment (https://www.ucl.ac.uk/isd/services/file-storage-sharing/data-safe-haven-dsh). Data and code supporting this manuscript are openly available via GitHub (https://github.com/hamishgibbs/huq_behaviour_clustering_paper).

## Acknowledgements

The authors acknowledge the following funding sources: ESRC UBEL Doctoral Training Partnership (ESRC: ES/P000592/1) (HG). Retail Business Datasafe (ESRC: ES/L011840/1) (JC). RME was also supported by the National Institute for Health Research (NIHR) Health Protection Research Unit (HPRU) in Modelling and Health Economics, a partnership between UK HSA, Imperial College London, and LSHTM (grant number NIHR200908). The views expressed are those of the authors and not necessarily those of the UK Department of Health and Social Care (DHSC), NIHR, or UKHSA.

## Notes

### Competing Interest Statement

The authors have declared no competing interest.

### Summary of Updates

This manuscript has been revised to focus on the role played by different mobile apps in defining the mobility dynamics reported by in-app GPS mobility data.

