## Supplemental Information for "Detecting behavioural bias in GPS location data collected by mobile applications"

### 1. In-app mobile phone location data

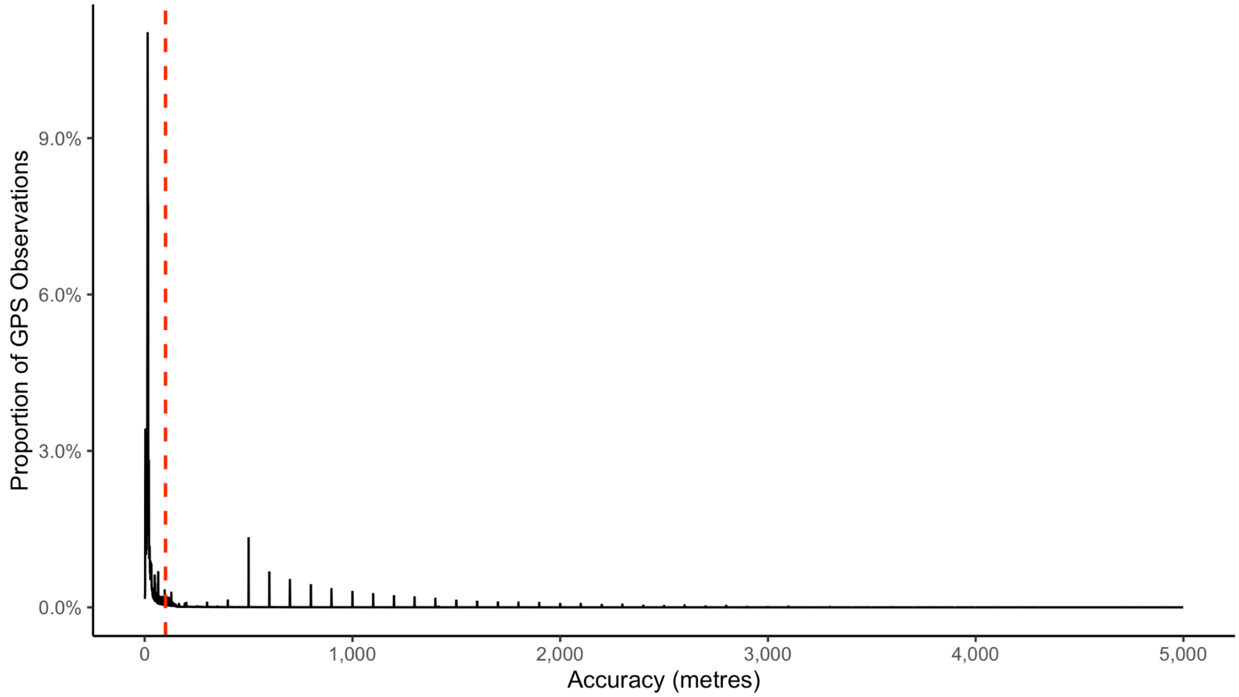


***Supplemental Figure 1.1. Threshold of accuracy values for GPS observations.*** *The distribution of position accuracy values for GPS observations from the original dataset. Values indicate a 68% confidence estimate that a device is within the given metres of the reported GPS coordinates. Higher values indicate lower confidence in position accuracy. Red dashed line shows the threshold defining "high accuracy" values (below 100m) used in this study.*

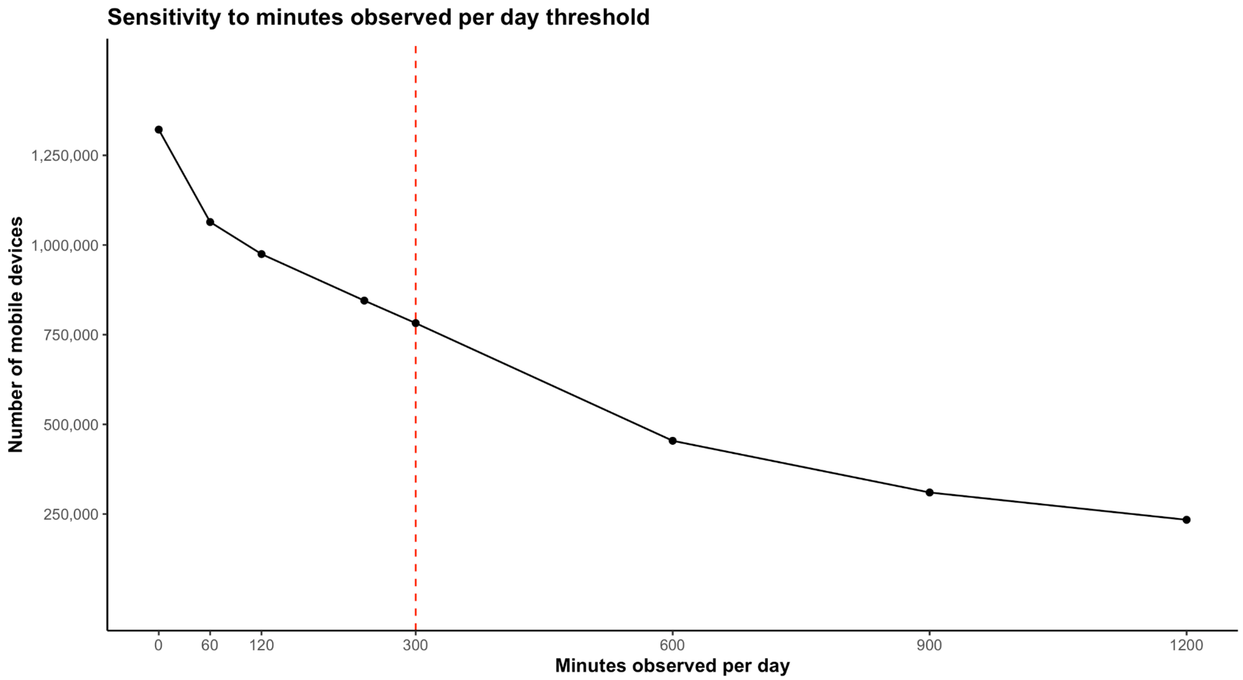


***Supplemental Figure 1.2. Sensitivity analysis of observation window threshold.*** *Sensitivity of sample selection to a minimum observation period. Red dashed line shows the 5 hour threshold used in this study.*

### 2. Principal component analysis

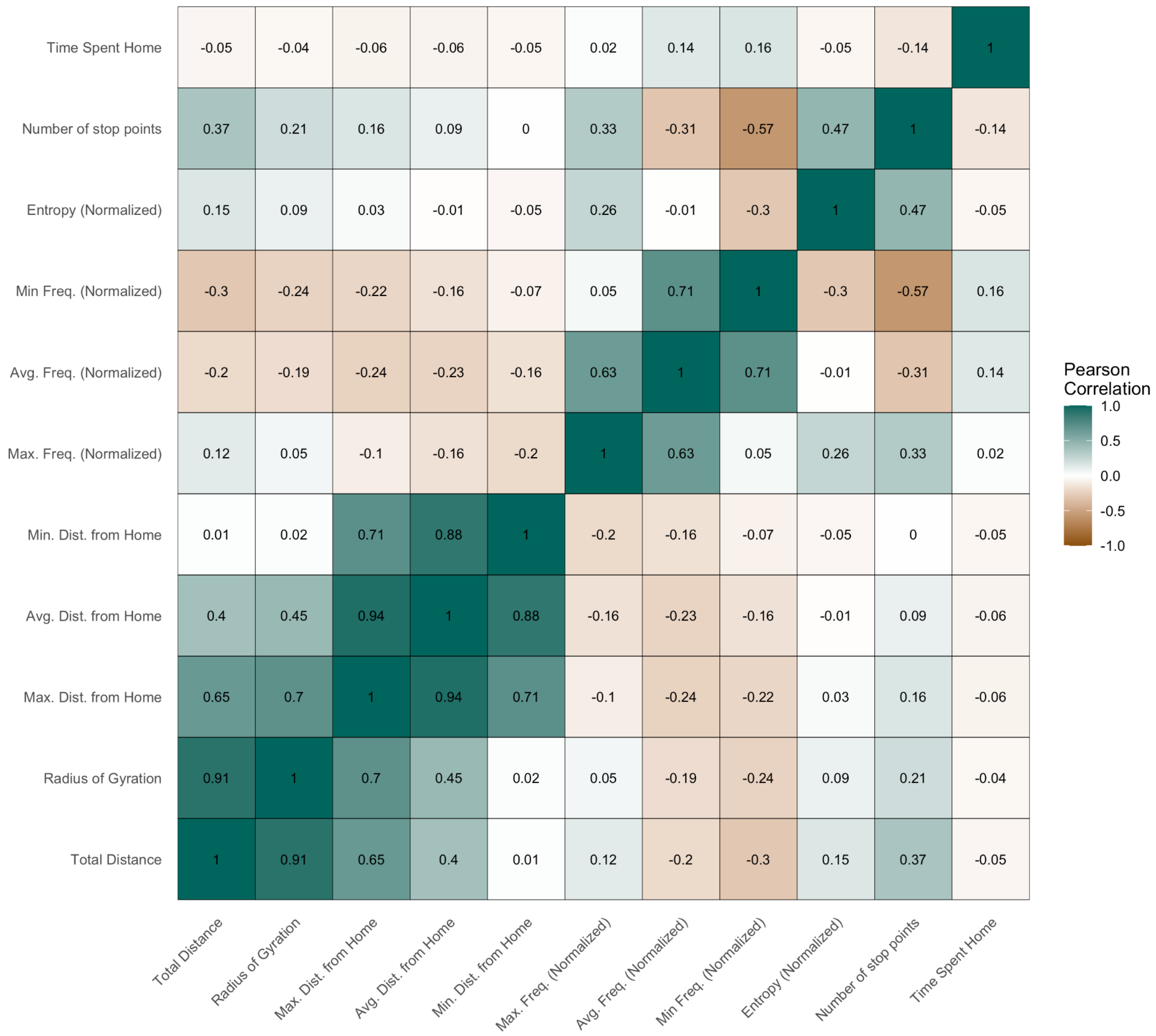


***Supplemental Figure 2.1. Correlation between input variables describing daily travel patterns.*** *Pearson correlation coefficient comparing measures of travel activity computed for each day.*

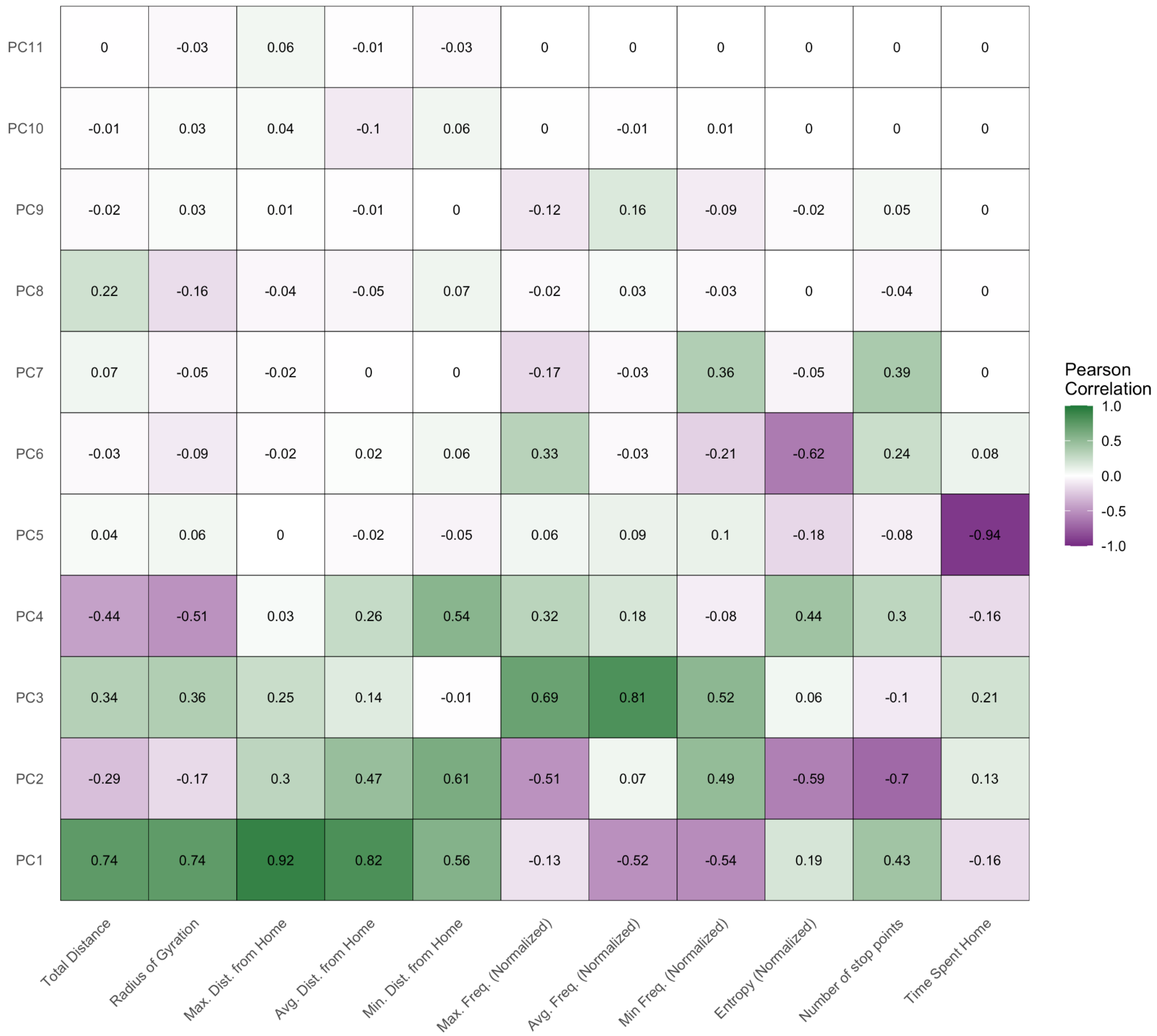


***Supplemental Figure 2.2. Correlation between input variables and principal components.*** *High correlation indicates greater contribution of an input variable to a specific principal component.*

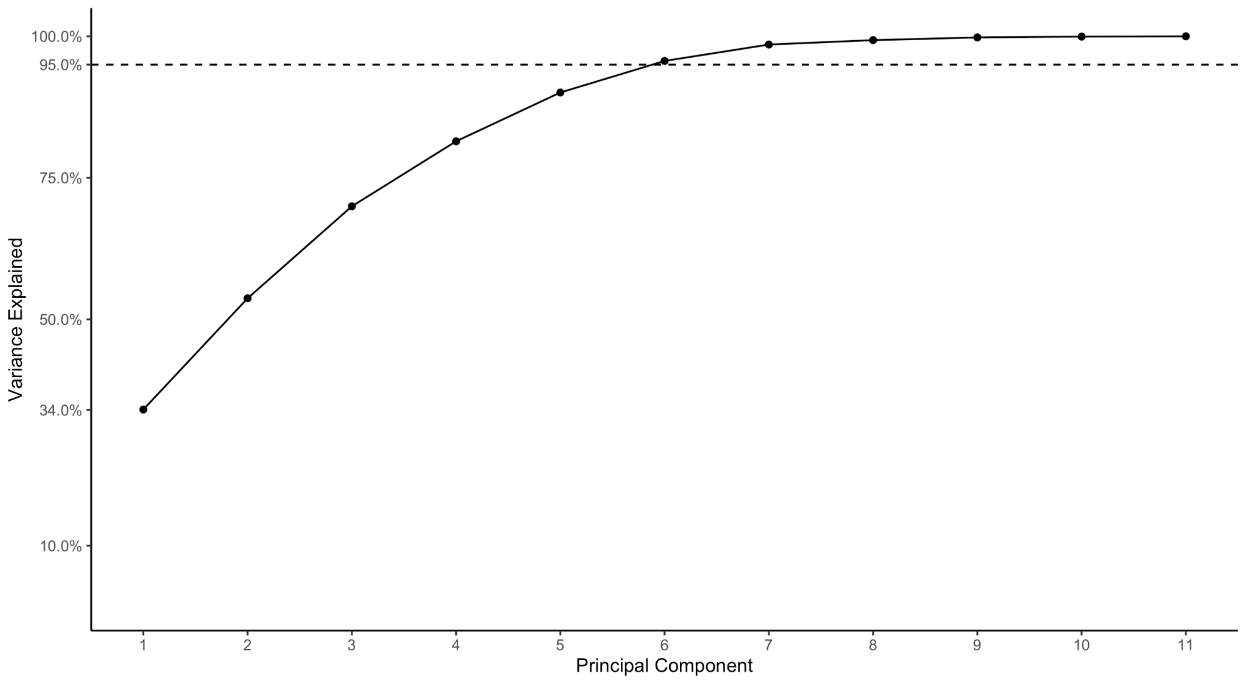


***Supplemental Figure 2.3. Variance explained by principal components (scree plot).*** *Dashed horizontal line indicates principal components capturing more than 95% of variance in the original dataset.*

### 3. Clustering

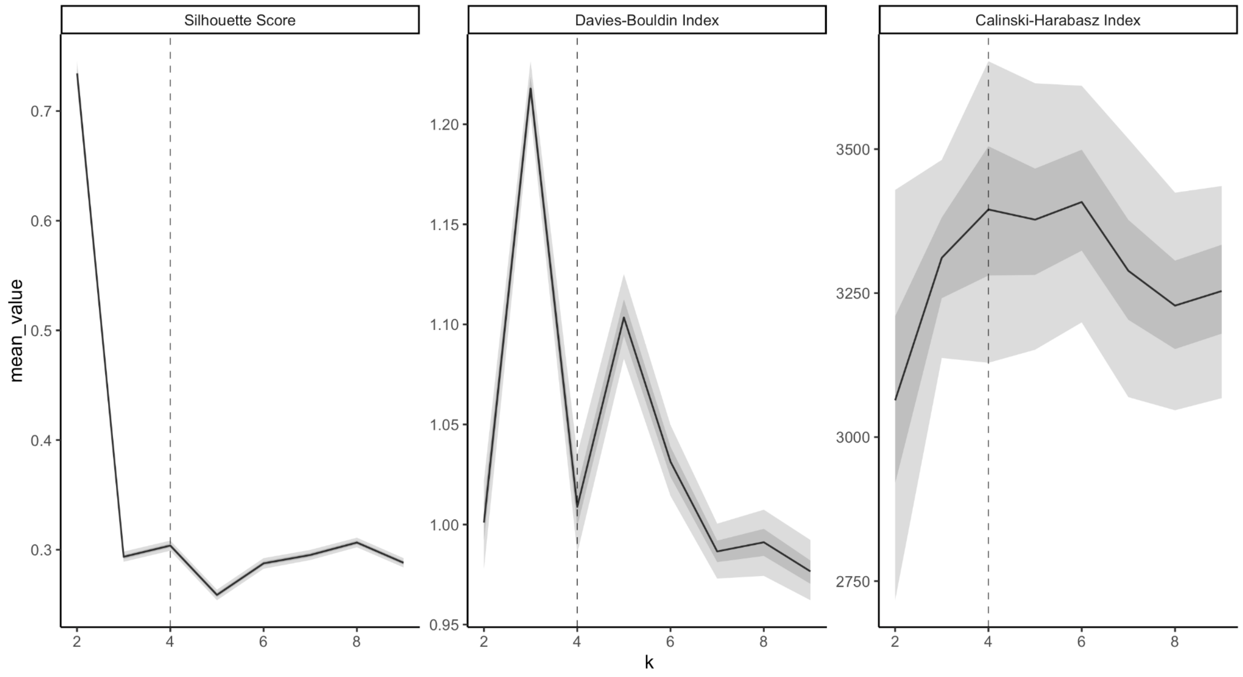

***Supplemental Figure 3.1. Quantitative cluster quality metrics.*** *Quantitative measures of cluster quality: Silhouette Score, Davies-Bouldin Index, and Calinski-Harabasz Index. Confidence intervals derived from bootstrapped estimates of cluster quality metrics (1,000 samples of 0.1% of the original dataset).*

### 4. Network Connectivity

| Resolution level | Average cell area ($km^2$) |
|---|---|
| 6 | 36.13 |
| 5 | 252.9 |
| 4 | 1,770.35 |
| 3 | 12,393.43 |

***Supplemental Table 4.1. H3 average cell area.*** *The average cell area of H3 hexagonal spatial grid cells (source: https://h3geo.org/docs/core-library/restable/).*

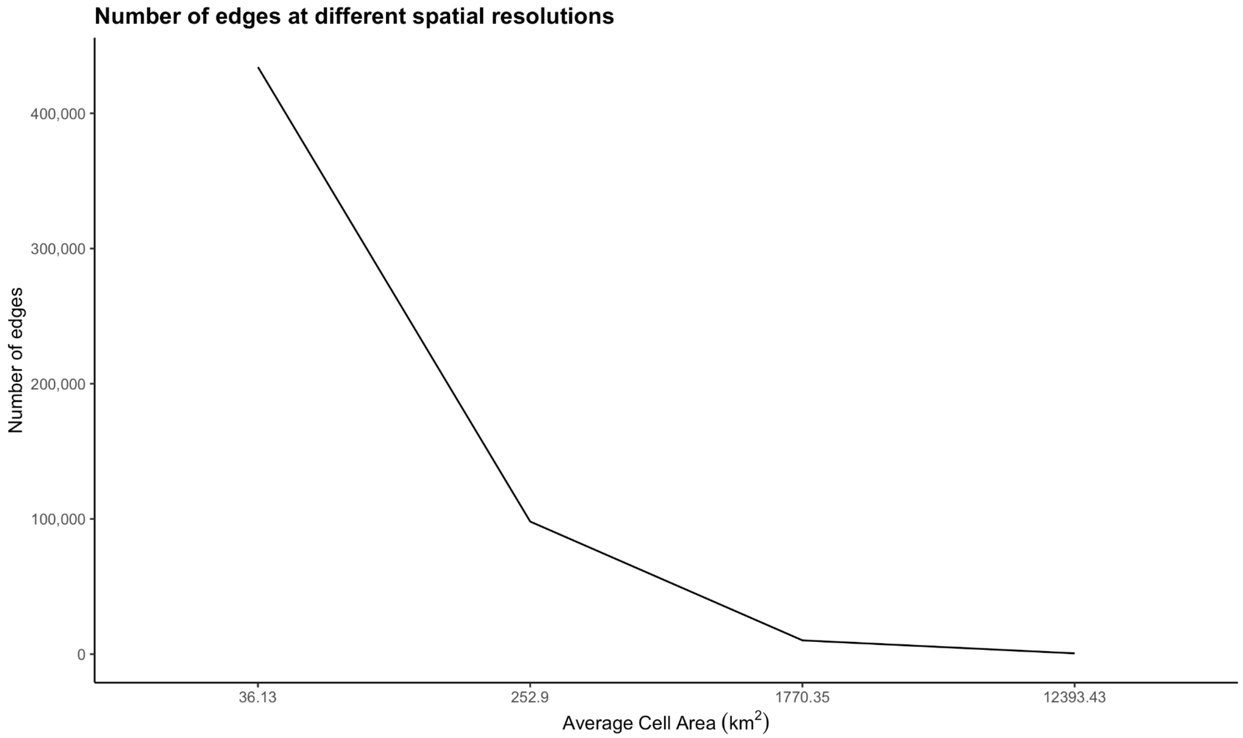


***Supplemental Figure 4.1. Number of edges in the travel network for varying sized grid cells.*** *Aggregation is based on a regular spatial grid (H3) defined by hierarchical spatial resolutions listed in Supplemental Table 4.1.*

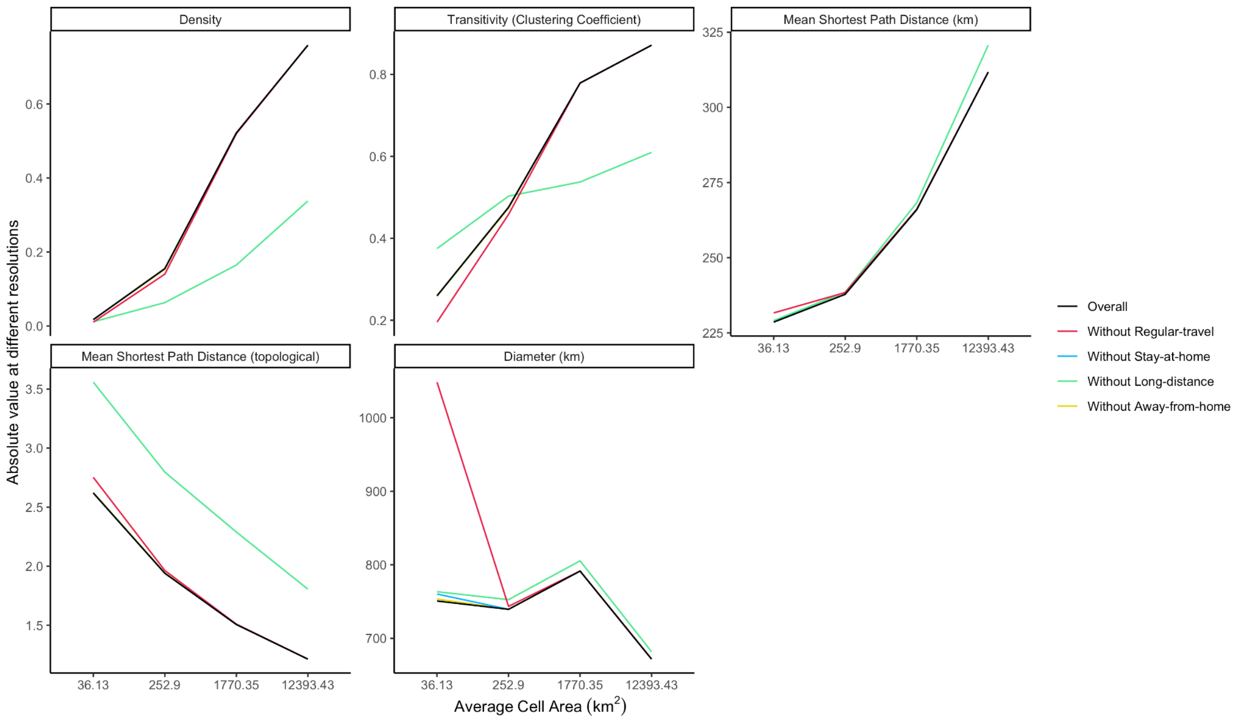


***Supplemental Figure 4.2. Network measures under different resolution spatial aggregations (absolute).*** *The effect of spatial aggregation on different measures of aggregated travel network characteristics, showing sensitivity of certain measures to spatial scale.*

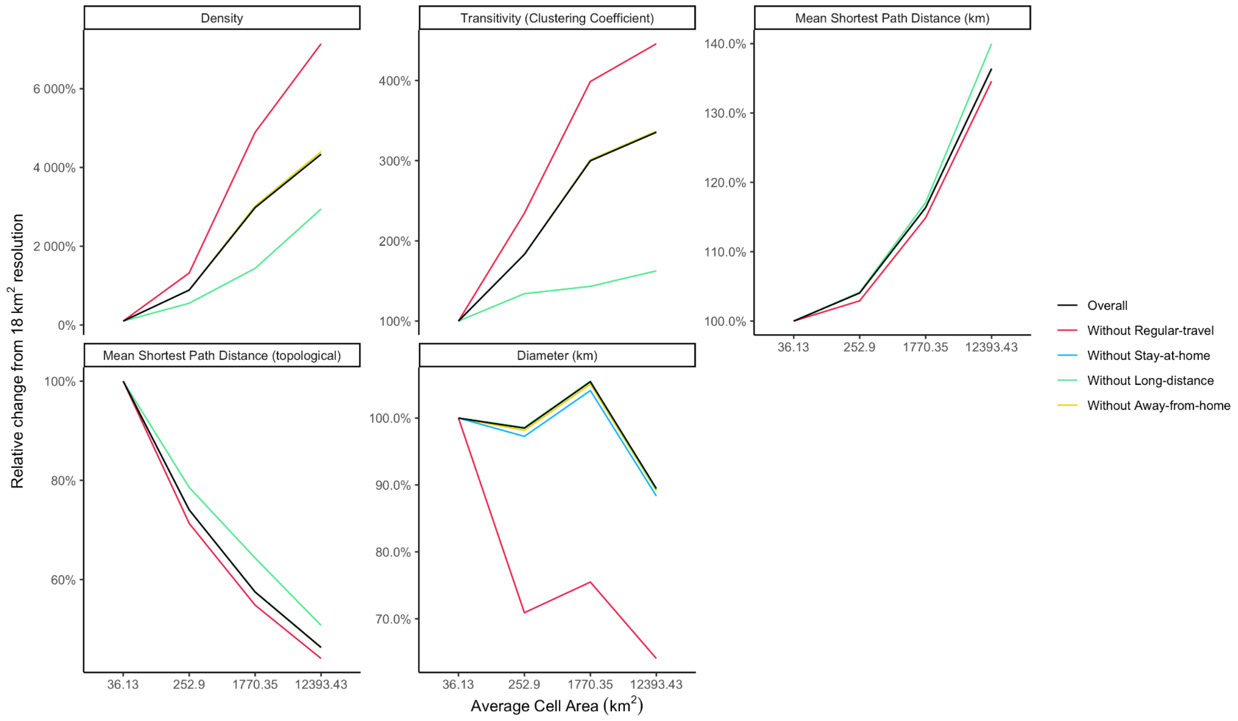

***Supplemental Figure 4.3. Network measures under different resolution spatial aggregations (relative).*** *The effect of spatial aggregation on different measures of aggregated travel network characteristics, calculated as a percent change from values at H3 resolution 6 (18.23 $km^2$ average cell area), showing sensitivity of certain measures to spatial scale.*
